# Mining Twitter to Assess the Determinants of Health Behavior towards Palliative Care in the United States

**DOI:** 10.1101/2020.03.26.20038372

**Authors:** Yunpeng Zhao, Hansi Zhang, Jinhai Huo, Yi Guo, Yonghui Wu, Mattia Prosperi, Jiang Bian

## Abstract

Palliative care is a specialized service with proven efficacy in improving patients’ quality-of-life. Nevertheless, lack of awareness and misunderstanding limits its adoption. Research is urgently needed to understand the determinants (e.g., knowledge) related to its adoption. Traditionally, these determinants are measured with questionnaires. In this study, we explored Twitter to reveal these determinants guided by the Integrated Behavioral Model. A secondary goal is to assess the feasibility of extracting user demographics from Twitter data—a significant shortcoming in existing studies that limits our ability to explore more fine-grained research questions (e.g., gender difference). Thus, we collected, preprocessed, and geocoded palliative care-related tweets from 2013 to 2019 and then built classifiers to:1) categorize tweets into promotional vs. consumer discussions, and 2) extract user gender. Using topic modeling, we explored whether the topics learned from tweets are comparable to responses of palliative care-related questions in the Health Information National Trends Survey.

## Introduction

As a relatively new but promising medical specialty created in 2007, palliative care adopts an interdisciplinary approach to provide physical, psychological, psychiatric, and spiritual support for terminally ill patients (e.g., advanced cancer patients) and their caregivers (e.g., family members). Palliative care is focused on relief from the symptoms and stress of a serious illness, with the goal is to improve the quality of life (QoL) for both the patients and their family. As a top priority identified to improve health care quality, palliative care has entered a period of rapid growth. However, due to misconceptions that palliative care is only for patients with unmanageable diseases, patients have been reluctant to accept this type of care; while oncologists also have delayed referring patients to specialized palliative care physicians for the same reason.^1^ According to a survey in 2011 commissioned by the Center to Advance Palliative Care (CAPC) with support from the American Cancer Society (ACS), nearly 70% people do not know about palliative care.^2^ This situation has not improved; as in a more recent 2016 survey study of 800 New York State residents, it was reported 73% of the respondents did not know what the term “*palliative care*” meant.^3^ However, after reading a brief description of palliative care services, 9 out of 10 survey respondents felt that palliative care is important for patients and their families.^3^ Our own study used the 2018 National Cancer Institute’s (NCI) Health Information National Trends Survey (HINTS) data—a nationally representative sample of U.S. adults—also confirmed that 71% had no knowledge of palliative care.^4^ Furthermore, even for those who felt confident in their knowledge of palliative care, around 15% respondents agreed that palliative care means giving up or stop other treatments, 30% respondents agreed that palliative care is the same as hospice care, and 40% respondents linked palliative care with automatically thinking of death.^4^ This lack of awareness and misconceptions in turn leads to underutilization of palliative care services. In fact, palliative services are underutilized among both cancer and noncancer populations. Data from the National Palliative Care Registry indicates that palliative care service penetration—the percentage of annual hospital admissions seen by the palliative care team—although has increased significantly since 2009, is still low at merely 4.8% as of 2015.^5^

To promote palliative care, the first critical step is to understand the factors that affect people’s health behavior towards palliative care. Recognized by the Integrated Behavior Model (IBM) as shown in ***Figure 1***, a general theory of behavioral prediction, individuals’ intention is the most important determinant of their health behaviors, while behavior intention is subsequently determined by attitude (e.g., feelings about the behavior), perceived norms (e.g., the social pressure one feels to perform the behavior), and personal agency (e.g., perceived control, self-efficacy).^6^ Other factors such as knowledge (i.e., skills to perform the behavior), environmental constraints (e.g., access to care), habits, and salience of the behavior can also directly affect individuals’ health behaviors. Traditionally, interviews, focus groups, and questionnaires are used to understand these determinants that affect individual’s behavior decision-making process. A few studies used these traditional approaches to examine the determinants of palliative care service utilization such as misconception, ignorance and lack of awareness of resources, and false hope.^7,8^ However, there are a number of shortcomings in traditional methods such as (1)low response rates and difficulties in recruitment, (2) high costs, (3) survey fatigue, (4) moderator bias, and (5) patients’ avoidances of sharing their real experience and feelings.^9^ Meanwhile, social media has brought rapid changes to the health communication landscape. Individuals are increasingly sharing a large amount of personal health information, including their health experience, on various social media platforms such as Twitter. These user-generated data provide unique insights into individuals’ health behaviors. On the other hand, researchers that use social media data are also facing methodological issues, such as the lack of sociodemographic information and how social media results can be compared with and/or supplement results from traditional survey studies.

**Figure 1.**
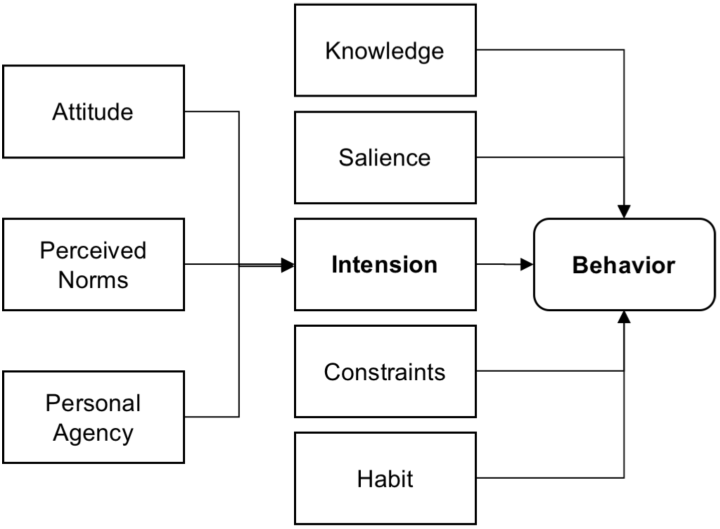
The integrated behavioral model.

In this study, we take the first step to fill these gaps aiming to (1) infer user attributes (i.e., age, gender, race) directly from user postings, and (2) compare social media findings with survey results guided by the IBM. In particular, we collected Twitter postings relevant to palliative care using a set of search keywords. We then built two machine learning models to classify these tweets into (1) relevant vs. irrelevant to palliative care discussions, and then within relevant tweets (2) promotional information (e.g., news, advertisements) vs. laypeople discussions. We used a topic modeling method to learn the themes of palliative care discussions on Twitter, extracted the user attributes (with an initial focus on age) and geolocation information of laypeople tweets, and assessed various associations between the learned Twitter topics and responses to palliative care-related questions in the 2018 NCI HINTS 5 Cycle 2 survey data^10^ to explore whether we can derived social media-based measures comparable to survey-based measures of palliative care. More specific, we aim to answer the following 3 research questions (RQs).

- **RQ1**. *What are the commonly discussed topics in promotional information and laypeople discussions on Twitter related to palliative care? Are consumers’ palliative care discussions on Twitter affected by promotional information?* (*i*.*e*., *through assessing the correlations between promotional palliative care-related information and consumers’ discussions in terms of topic distributions*)
- **RQ2**. *Can the learned topics be mapped to the constructs in the IBM? If so, are the geographic distributions of the learned topics comparable to the determinants measured from HINTS survey?*
- **RQ3**. *Can we extract a user’s gender from their Twitter postings? If so, are the geographic distributions of the learned topics comparable to the determinants measured from HINTS survey stratified by gender?*

If successful, the learned topics can potentially be used as measures of IBM constructs that ultimately can help us identify and understand the determinants of consumers’ health behaviors towards palliative care.

## Methods

The general process of our approach, as shown in ***Figure 2***, is to (1) collect palliative care-related tweets, (2) classify tweets into consumers’ discussions and promotional information, (3) predict Twitter users’ gender based on their posts, (4) abstract topics from both consumers’ discussions and promotional information, and (5) answer our 3 *research questions based on the topic distributions*.

**Figure 2.**
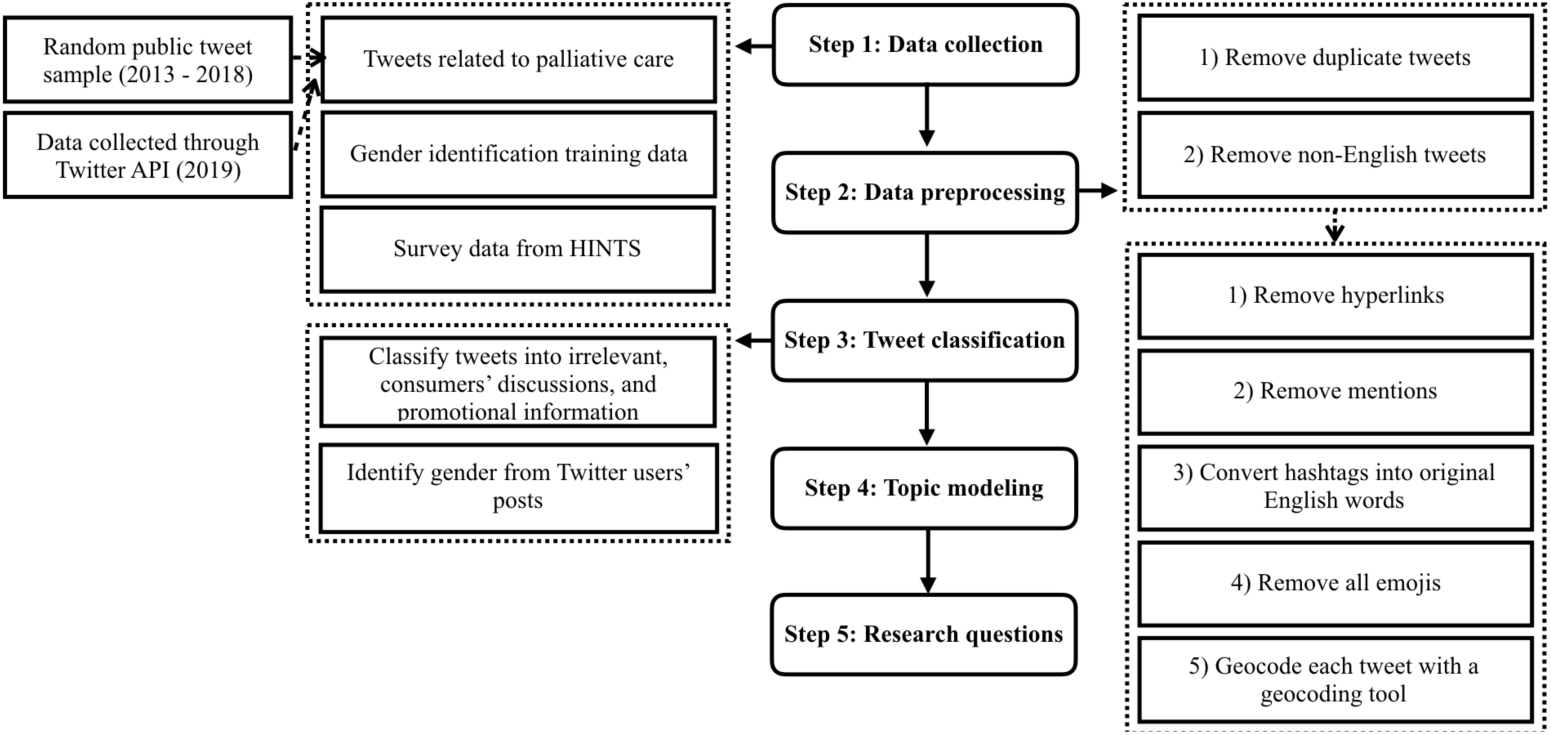
The general analysis workflow.

### Step 1: Data collection

#### Tweets related to palliative care

We collected two Twitter datasets related to palliative care: (1) we collected 1,110,632 palliative care-related tweets from March 1, 2019 to May 20, 2019 using a crawler that we developed previously,^11,12^ based on a set of palliative care-related keywords (e.g., “*palliative care*” and “*palliative medicine*”). We generated the keywords using a snowball sampling process: started with a set of seed keywords, then searched on Twitter with these keywords to retrieve a sample of tweets, evaluated the relevance of each tweet, and identified new keywords. We did this process iteratively until no new keywords were found; and (2) we used the keywords developed above and identified 333,888 related tweets on a database of public random tweets (i.e., from 2013 to 2017, which we collected using the same Twitter crawler).

#### Training data for gender identification

To develop classifiers to identify gender, we obtained the author profiling task dataset from PAN 2018, which focused on gender identification with Twitter data.^13^ A set of 100 tweets and 10 images were provided for each user and the users were grouped by three languages including English, Arabic and Spanish. In this study, we focused on English tweets and text data only. The PAN 2018 dataset originally provided 3,000 Twitter users for training and 1,900 Twitter users for testing, where the truth labels were released at the end of the competition. Thus, in total, we obtained 4,900 labeled Twitter users from the PAN 2018 dataset and the gender distribution balanced (i.e., 2,450 male and 2,450 female) without missing labels, tweets, and images.

#### Survey data from HINTS

Further, we obtained survey data from 2018 HINTS 5, Cycle 2.^10^ HINTS is a nationally representative survey on public’s use of cancer and health-related information. We extracted responses from 3,504 respondents who answered 12 palliative care-related questions. We also obtained state-level geographic information and full-sample weight (i.e., to calculate population estimates) of each respondent.

### Step 2: Twitter data preprocessing

We preprocessed the Twitter data to eliminate duplicate tweets and tweets that were not written in English. To develop the tweet classifiers, we further preprocessed the tweets following the steps used by GloVe ^14^ : (1) removed hyperlinks, (2) removed mentions, (3) converted hashtags into English words (e.g., convert “#palliativecare” to “*palliativecare*”), (4) removed all emojis, and (5) geotagged each tweet to a US state with a tool we developed previously.^15^ Twitter has three options to attach geographic information to each tweet or a user profile: (1) a geocode (i.e., latitude and longitude) or a geographic ‘*place’* can be attached to each tweet, if it is posted with a GPS-enabled mobile device or the user chose to tag it with a ‘*place’*; (2) the associated user profile can be geocoded (either to a GPS location or a ‘*place’*); and (3) the free-text ‘*location’* attribute can be filled by the user. If geocodes were available, we attempted to resolve the locations through reverse geocoding using GeoNames,^**16**^ a public geographical database. Only 0.91% of tweets and 0.45% of Twitter user profiles in our datasets have geocodes (i.e., latitude and longitude pairs). For most tweets, we matched the free-text ‘*location’* with lexical patterns indicating the location of the user such as a state name (e.g., “*Florida*”) or a city name in various possible combinations and formats (e.g., *“——, fl*”). For topic modeling, we also lemmatized each word and removed stop words (e.g., “*it*”, “*is*”).

### Step 3: Tweet classification

Even though a tweet contains keywords related to palliative care, the tweet may not be relevant to the palliative care discussions (e.g., *“Comfort Care Toilet Paper”*). Further, to answer RQ1, we need to separate promotional information on Twitter from genuine consumer discussions. Thus, we developed a two-step process with two classification models to categorize the massive number of tweets into 3 groups (i.e., irrelevant, promotional, and consumer discussions).

To develop the classification models, we randomly selected and annotated 1,839 tweets from the overall dataset based on keyword distributions to create a training set. We then experimented with one machine learning (i.e., Random Forest [RF]) and two deep learning algorithms (i.e., Convolutional Neural Networks [CNN) and Long Short-Term Memory [LSTM)) to categorize the tweets into the 3 groups. We choose RF because it has good out-of-box performance for Twitter text classification tasks,^17^ while deep learning methods have shown state-of-art performance surpassing traditional methods.^18^ We implemented the RF via the scikit-learn^19^ library and used n-grams to convert tweet text into feature vectors. We implemented the CNN and LSTM in Keras^20^ on the top of the Tensorflow^21^ framework and used the GloVe^14^ pretrained 100 dimension Twitter word embeddings in the convolutional layer.

Demographic data are often missing or restricted to access in many social media platforms. For example, Twitter users are not required to provide detailed demographics (e.g., race) or some of the attributes are restricted to only visible to “friends” (e.g., year of birth). The missing of such information makes it challenging to answer more specific research questions across subpopulation groups (e.g., “*Is there any gender difference in terms of individual’s attitude towards palliative care?*”). However, new machine learning (ML) research can infer users’ demographic attributes with high accuracy, including age,^22^ gender,^23^ and race/ethnicity. In this study, we leveraged the PAN 2018 author profiling task dataset that focused on gender identification of Twitter users, whereas both text (i.e., 100 tweets) and image (i.e., 10) were available. We focused on the text data when building our classifiers. Following the same approach above, we experimented with three classification algorithms (i.e., RF, CNN, and LSTM). 80% of the 4,900 Twitter users were used for training and the rest as an independent testing data.

### Step 4: Topic modeling

Topic modeling is an unsupervised approach that can extract themes/topics from a collection of documents. We used the biterm topic model (BTM) to identify the latent topics from all palliative care-related tweets (i.e., combined both promotional information and consumers’ discussions).^**24**^ Different from the commonly used latent Dirichlet allocation (LDA) model using document-level word co-occurrence patterns to reveal topics, BTM directly models the generation of word co-occurrence patterns (i.e., biterms) in the whole corpus. Doing so, BTM addresses the data sparsity issue commonly found in short documents, and thus, outperforms traditional topic models like LDA on tweets.^**24**^ Similar to LDA, BTM represents each document (i.e., a tweet in our case) as a mixture of latent topics, and each topic can generate words with certain probabilities. Although topic modeling is unsupervised, the number of topics needs to be set a priori as a parameter. In previous studies,^12^ we tested various statistical measures (e.g., KL divergence) for determining the optimal number of topics and found that these metrics often do not converge and the number of topics discovered does not conform to human judgments. Additional qualitative analysis of the generated topics to determine their quality is still necessary. Learned from this experience, we picked a relatively large number (i.e., 50), so that the learned topics would cover all potential discussion themes. To evaluate the quality of the learned topics, two annotators were presented with the word clouds and the top 20 tweets associated the topics and were asked to assign each topic a label based on their judgments, independently. Each annotator was also asked to assess the quality of each topic. A topic is considered of low quality if (1) the keywords in the topic did not represent a cohesive concept, or (2) more than half of the 20 tweets were assigned to the topic incorrectly (i.e., the content of an assigned tweet does not reflect the keywords in the topic’s word cloud).

The nature of the BTM allows all topics to occur in the same tweet with different probabilities, while topics with low probabilities might not actually exist. Thus, we needed to determine a cutoff probability value to select the most representative and adequate topics to be assigned to each tweet. We tested a range of cutoff values and manually evaluated a random sample of tweets (i.e., 100) for each tested cutoff value to determine whether the topics (whose probabilities were larger than the cutoff) assigned to each tweet were correct. We then selected the lowest cutoff where more than 70% of topic assignments were adequate. Note that the cutoff values for promotional vs. consumer tweets were established separately.

### Step 5: Answer the three research questions

We answered the 3 RQs through analyzing the discussion themes learned from the BTM in the following steps. For **RQ1** (i.e., “*What are the commonly discussed topics in promotional information and laypeople discussions on Twitter related to palliative care? Are consumers’ palliative care discussions on Twitter affected by promotional information?*”), we visualized the learned latent topics as word clouds and qualitatively analyzed these topics. We identified the top 10 topics within in each of the two categories of tweets (i.e., promotional information vs. consumer discussions) and examined the common topics across the two. We plotted the monthly trends of the topics for both categories and then calculated the Pearson correlation coefficient between the two in terms of monthly tweet volumes for each topic to assess whether consumer discussions are impacted by promotional information.

For **RQ2** (i.e., “*Can the learned topics be mapped to the constructs in the IBM? If so, are the geographic distributions of the learned topics comparable to the determinants measured from HINTS survey?*”), we first mapped high-quality topics to IBM constructs through manually examining each topic’s word-cloud and a sample of 20 associated consumer tweets by two annotators (HZ and YP). For example, “*fundraising for palliative care*” with a sample consumer tweet—“*I am fundraising for Springhill Hospice* (*Rochdale*). *Please donate at my JustGiving page*”—can be mapped to the “*environmental constraint*” construct in IBM. A topic is excluded if it does not represent consumer discussions (i.e., more than 10 out of the 20 sample tweets are irrelevant to the topic theme). We then grouped the 12 palliative care-related HINTS questions into question groups (QGs) and mapped the QGs to IBM constructs. For example, we merged “*Imagine you had a strong need to get information about palliative care. Where would you go first to get information”* and *“Image you had a strong need to get information about palliative care. Which of the following would you most trust as a source of information about palliative care?”* in to a *“palliative care trust information”* group. We then established the correlations between consumer-related palliative care Twitter topics and population estimates derived from the palliative care-related HINTS questions at state level. Note that we excluded promotional information because (1) many of the user accounts are organizations and public figures, and (2) these promotional posts do not represent consumer thoughts about palliative care thus does not reflect consumer’s behavioral determinants towards palliative care. To calculate the correlations, we first calculated normalized geographic distribution of topic discussion rates (i.e., Twitter users who discussed a specific topic divided by the total number of Twitter users who posted palliative care-related tweets in a state). From survey data, to obtain the normalized geographic distribution of HINTS response rates, we divided the number of respondents with the answers of interest (e.g., responded *“strongly disagree”* to *“Accepting palliative care means giving up”*) by the total number of respondents for each state considering each respondent’s full sample weight in HINTS. Considering that we grouped questions into QGs, we also combined answers for all questions in that QG (i.e., if a respondent responds with the interested answer for any question in that QG). After normalized both Twitter and survey data, we calculated the Spearman’s rank correlations between Twitter topics and the population estimates (derived from HINTS survey responses to each QG) in terms of their geographic distributions.

For **RQ3** (i.e., “*Can we extract user attributes with an initial focus on gender from laypeople’s Twitter postings? If so, are the geographic distributions of the learned topics comparable to the determinants measured from HINTS survey stratified by gender?*”), we used the gender identification classifier developed in **Step 3** to extract the gender information of each Twitter user whose tweets were classified as consumer discussions based on their most recent 100 tweets (regardless of whether the tweets are related to palliative care or not). Similar to the approach for **RQ2**, we then calculated the Spearman’s rank correlations between Twitter topics and the population estimates (derived from HINTS survey responses to each QG) in terms of their geographic distributions but stratified by gender.

## Results

### Twitter datasets

The snowballing process generated 77 keywords related to palliative care. Based on these 77 keywords, our palliative care Twitter data were collected from two different sources: (1) we collected 1,110,632 tweets related to palliative care from March 1, 2019 to May 20, 2019 (81 days). After removing duplicates, non-English tweets, and tweets without keywords (e.g., *“@Palliative_Bio”*, where the keyword “palliative” is part of a user name), 333,888 tweets remained; and (2) we used the same list of keywords to extract tweets from our historical random sample database, which was collected using the Twitter steaming application interface (API), from January 1, 2013 to May 31, 2018. From this dataset, we collected a total of 213,559 tweets. After removing duplicates, non-English tweets, and tweets without keywords, 95,084 tweets were left. ***Table 1*** shows the number of tweets by year.

**Table 1.** Number of tweets by year from the two data sources

| Year | # of tweets | # after preprocessing<sup>a</sup> | # of geo-tagged tweets | # of users | # of geo-tagged user |
| --- | --- | --- | --- | --- | --- |
| 2013 | 23,764 | 12,355 | 2,438 | 9,364 | 1,603 |
| 2014 | 29,258 | 14,282 | 2,958 | 10,583 | 1,985 |
| 2015 | 35,874 | 15,323 | 3,496 | 10,917 | 2,284 |
| 2016 | 47,767 | 20,399 | 4,425 | 13,264 | 2,747 |
| 2017 | 48,119 | 20,833 | 4,515 | 14,296 | 2,818 |
| 2018 | 28,777 | 11,892 | 2,218 | 9,446 | 1,698 |
| 2019 | 1,110,632 | 333,888 | 73,640 | 205,679 | 44,194 |
| **Total** | **1,324,191** | **428,972** | **93,690** | **273,549** | **57,329** |

<sup>a</sup>Preprocessing: removed 1) duplicates 2) non-English, and 3) tweets without keywords

### Tweets classification and identification of user gender

We explored 3 classification algorithms (i.e. RF, CNN and LSTM) to category the tweets into 3 categories (i.e., irrelevant, promotional information, and consumer discussions) using a two-step process. We used 80% of the annotated data for training and the performance was measured on the reserved 20% independent testing data. As shown in ***Table 2***, the CNN models outperformed other algorithms in both tasks (i.e., (1) relevant vs. irrelevant; and subsequently (2) promotional vs. consumer within relevant tweets). Thus, we adopted the CNN models as the final classifiers. The CNN classifier identified 371,880 relevant tweets (out of 428,972 tweets with palliative care-related keywords). Out of the 371,880 relevant tweets, 258,284 tweets were classified as promotional information; and 113,596 tweets (82,172 Twitter users) were classified as consumer discussions. Within consumer tweets, 29,993 tweets (27,089 Twitter users) can be geotagged to a US state.

**Table 2.** Performance of tweet classification and gender identification models

| Model | Precision | Recall | F-score |
| --- | --- | --- | --- |
| **Relevant vs. Irrelevant (1,397 vs. 442)** |  |  |  |
| RF | 0.73 | 0.66 | 0.68 |
| CNN | **0.75** | **0.76** | **0.76** |
| LSTM | 0.75 | 0.77 | 0.74 |
| **Promotional information vs. Consumers' discussions (1,082 vs. 315)** |  |  |  |
| RF | 0.76 | 0.75 | 0.76 |
| CNN | **0.78** | **0.79** | **0.79** |
| LSTM | 0.77 | 0.76 | 0.76 |
| **Gender identification (male: 2,450 vs. female: 2,450)** |  |  |  |
| RF | 0.65 | 0.64 | 0.64 |
| CNN | **0.80** | **0.80** | **0.80** |
| LSTM | 0.58 | 0.58 | 0.57 |

We explored the same 3 classification algorithms (i.e., RF, CNN and LSTM) to develop a gender classifier based on Twitter users’ tweets. The PAN 2018 dataset provided 100 tweets for each user. For RF, we used the n-grams scheme to extract features from text. For deep learning models (i.e., CNN and LSTM), we used pretrained GloVe Twitter word embeddings to convert tweets into feature vectors. We trained each model on the 3,000 users training set and measured performance on the reserved 1,900 Twitter users testing dataset. ***Table 2*** shows the performance of the gender classification models; and CNN again outperformed other models. The CNN classifier identified 6,322 males and 20,787 females from the 27,089 lay consumers who discussed palliative care on Twitter.

### Topic modeling

We trained a BTM using all the 371,880 tweets (i.e., both promotional and consumer tweets) relevant to palliative care with the number of topics set at 50 to extract as many topics as possible. Two annotators (i.e., kappa: 0.79) reviewed the 50 topics and a sample of 20 tweets associated with topic to (1) assess topic quality and (2) merge topics with similar themes. Even through the BTM was trained using both promotional and consumer tweets, some of topics might not be more prevalent in consumer discussions or vice versa in promotional tweets. Thus, a high-quality topic identified based on consumer tweets might not be of high quality in terms of promotional tweets. Through this process, we identified 17 high-quality topics from consumer tweets and 12 high-quality topics from promotional tweets.

We determined the cutoff probabilities for consumer and promotional tweets separately. For consumer tweets, the cutoff probability is 0.2, where 72% of the 100 randomly selected consumer tweets’ topic assignments were adequate. For promotional tweets, the cutoff value is 0.3, where 70% of the selected tweets had adequate topic assignments. We were able to assign topics to 91.4% (i.e., 27,426) of the 29,993geocoded consumer tweets.

### Research questions

#### RQ1: What are the commonly discussed topics in promotional information and consumer discussions on Twitter related to Palliative care? Are consumer discussions affected by promotional information?

As shown in ***Table 3***, we identified top 10 discussed topics across consumer and promotional tweets. ***Figure 3*** shows the word clouds and sample tweets of the top 3 topics in consumer discussions and promotional information..

**Table 3.** Top 10 most discussed topics in consumer discussions and promotional information related to palliative care

| ID | Topic in Consumer | Consumer Tweets (%)<br>N = 113,596 | ID | Topic in Promotional | Promotional Tweets (%)<br>N = 258,284 |
| --- | --- | --- | --- | --- | --- |
| 31, 45 | Family member in hospice | 26,866 (23.7 %) | 6 | Fundraising | 16,713 (6.5%) |
| 38 | Care for non-viable newborn | 8,692 (7.7%) | 17 | Support for hospice | 12,320 (4.8%) |
| 6 | Fundraising | 4,143 (3.6%) | 33 | Palliative care events | 7,647 (3.0%) |
| 25 | Palliative care for newborn with bilateral renal agenesis | 3,718 (3.3%) | 15 | Debate about palliative care is infanticide | 5,872 (2.3%) |
| 41 | Documentary of patients | 2,924 (2.6%) | 38 | Care for non-viable newborn | 3,942 (1.5%) |
| 35 | End of life care | 2,557 (2.3%) | 45 | Family members in hospice | 3,119 (1.2%) |
| 17 | Support for hospice | 2,312 (2.0%) | 3 | A sexually assaulting incident in hospice | 2,670 (1.0%) |
| 1 | Comfort care for unwanted newborn | 2,266 (2.0%) | 39 | People entered hospice care | 2,485 (1.0%) |
| 11 | Hospice patient and people against Trump policy | 2,128 (1.9%) | 5 | Pioneers in palliative care and palliative medicine | 2,210 (0.9%) |
| 15 | Debate about palliative care is infanticide | 2,120 (1.9%) | 11 | Hospice patient and people against Trump policy | 1,420 (0.5%) |

**Figure 3.**
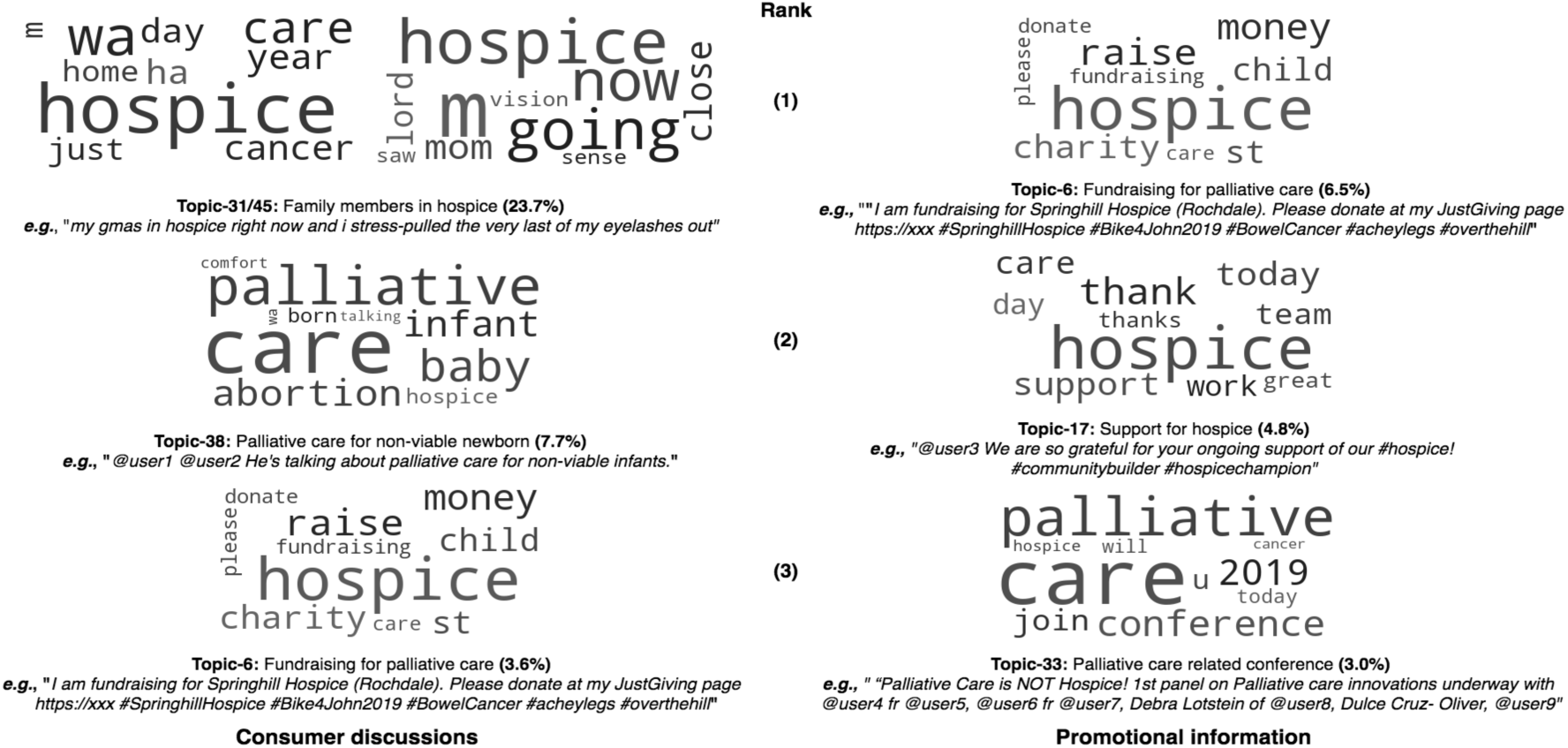
Word clouds of top 3 topics from consumer discussions and promotional information.

There are 12 high-quality topics in both promotional and consumer tweets as shown in ***Table 4***. 5 out of the 17 high-quality topics are unique to consumer discussions of palliative care (e.g., topic 14: “*people ask for help and pray for hospice patients*” and topic 21: “*People talked about their friends in hospice*”). 7 out of the 12 common topics are correlated between promotional and consumer tweets (p < 0.05) in terms of their monthly volumes.

**Table 4.** Common topics across promotional and consumer tweets and their correlations

| Topic ID | Common Topic across Promotional and Consumer Tweets | Correlation Coefficient |
| --- | --- | --- |
| 1 | Comfort care for unwanted newborn | 0.34 ($p < 0.01$) |
| 15 | Debate about palliative care is infanticide | 0.02 ($p = 0.88$) |
| 25 | Palliative care for newborn with bilateral renal agenesis | 0.98 ($p < 0.01$) |
| 38 | Palliative care for non-viable newborn | 0.98 ($p < 0.01$) |
| 5 | Pioneers in palliative care and palliative medicine | 0.69 ($p < 0.01$) |
| 6 | Fundraising for palliative care | 0.98 ($p < 0.01$) |
| 31, 45 | Family members in hospice | 0.35 ($p < 0.01$) |
| 33 | Palliative care related events | 0.98 ($p < 0.01$) |
| 35 | End of life care | 0.98 ($p < 0.01$) |
| 41 | Documentary of hospice patients | 0.01 ($p = 0.91$) |
| 3 | A hospice patient's final request | 0.07 ($p = 0.56$) |
| 17 | Support for hospice | 0.99 ($p < 0.01$) |

#### RQ2: Can the learned topics be mapped to the constructs in the IBM? If so, are the geographic distributions of the learned topics comparable to the determinants measured from HINTS survey?

16 out of the 17 high-quality consumer topics were mapped to 4 different IBM constructs: (1) knowledge (1 topic, e.g., topic 33: “*Palliative care related events*”), (2) attitude (9 topics, e.g., topic 1: “*comfort care for unwanted newborn*” and topic 15: “*debate on palliative care is infanticide*”), (3) perceived norm (5 topics, e.g., topic 31/45: “*family member in hospice*”), and (4) constrains (1 topic, e.g., “*fundraising*”).

We grouped 12 palliative care-related HINTS questions into 11 QGs and mapped the 11 QGs to 4 types of IBM constructs: (1) knowledge (2 QGs), (2) attitude (6 QGs), (3) perceived norm (2 QGs), and (4) habit (1 QGs). We then explored Spearman’s rank correlations between the geographic distributions of the discussion rates (i.e., the proportion of Twitter users who discussed that topic) for each of the 17 high-quality consumer topics in Twitter and the response rates (i.e., the proportion of people who gave the answer of interest) for each of the palliative care-related QGs in HINTS. ***Table 5*** shows examples of (1) the mappings of Twitter topics in consumer tweets to corresponding IBM constructs, and (2) correlations between Twitter topics and responses in HINTS.

**Table 5.** Examples of mapping topics in consumer discussions to the palliative care-related survey questions in HINTS and corresponding constructs in the Integrated Behavior Model (IBM).

| Palliative care-related survey questions in HINTS | IBM Construct | Correlation with the 17 high-quality consumer topics |  |  |
| --- | --- | --- | --- | --- |
|  |  | Both gender | Female | Male |
| QG1.a How would you describe your level of knowledge about palliative care?<br>*Answer:* I’ve never heard of it | Knowledge | Topic 5 - “pioneers in palliative care and palliative medicine” ($\rho$: -0.47; $p < 0.01$) | Topic 21 - “people talked about their friends in hospice” ($\rho$: -0.37; $p = 0.03$) | Topic 3 - “A sexually assaulting incident in hospice” ($\rho$: 0.41; $p < 0.02$) |
| QG2.b To me, the goal of palliative care is to help friends and family to cope with a patient’s illness<br>*Answer:* Strongly/Somewhat disagree | Attitude | Topic 31/45- “family members in hospice” ($\rho$: -0.39; $p = 0.02$) | Topic 31/45- “family members in hospice” ($\rho$: -0.38; $p = 0.03$) | Topic 35 - “End of life care” ($\rho$: 0.39; $p = 0.03$) |
| Q7.a It is a doctor's obligation to inform all patients with cancer about the option of palliative care<br>**Answer:** Strongly/Somewhat disagree | Perceived Norm | Topic 33 - "palliative care related events" ($\rho$: 0.38; $p$ = 0.03) | N/A | Topic 35 - "End of life care" ($\rho$: 0.36; $p$ = 0.04) |
| Q11.b Imagine you had a strong need to get information about palliative care.<br>1) Where would you go first to get information?<br>2) Which of the following would you most trust as a source of information about palliative care?<br>**Answer:** Health care provider (doctor, nurse, social worker) | Habit | Topic 6 - "Fundraising for palliative care" ($\rho$: 0.36; $p$ = 0.04) | N/A | NA |

#### RQ3. Can we extract a Twitter user’s gender from users’ Twitter postings? If so, are the geographic distributions of the learned topics comparable to the determinants measured from HINTS survey stratified by gender?

The gender classifier identified 6,322 males and 20,787 females from the 27,089 Twitter users whose tweets were classified as consumer discussions. Similarly, we then explored Spearman’s rank correlations between the geographic distributions of the discussion rates in Twitter and the response rates in HINTS stratified by gender shown in ***Table 5***.

## Discussion and conclusion

In this study, guided by the integrated behavioral model, we explored the potential of using user-generated content on Twitter to assess the determinants of consumers’ behaviors towards palliative care, which have been traditionally measured through surveys. A secondary goal is to assess the feasibility of extracting user demographics from Twitter data—a significant shortcoming in existing studies that limits our ability to explore more fine-grained research questions (e.g., gender difference). We collected palliative care-related tweets (i.e., 2013 to 2019), built classifiers to 1) categorize tweets into promotional vs. consumer discussions, and 2) extract user gender directly from user postings, applied topic modeling to abstract themes of consumer discussions, and subsequently answer 3 research questions. For RQ1, we found 4 out of the 12 common topics in both consumer and promotional tweets are related to newborns: (1) topic 1:*“Comfort care for unwanted newborn”*, (2) topic 15:*“debate about palliative care is infanticide”*, (3) topic 25: *“palliative care for newborn with bilateral renal agenesis”*, and (4) topic 38:*“Palliative care for non-viable newborn”*. One of the most difficult ethical dilemmas for parents and pediatric physicians arises when a child has complex chronic conditions that may not be curable and cause discomfort with no prospect of any improvement on quality of life. In the context of medical futility, it is harmful to prolong medical treatment. Then, palliative care enters into the discussions of parents and physicians when the primary goal of treatments is no longer focused on curing a condition but on making your baby as comfortable as possible^25^. In a rally in Panama City Beach, Florida on May 8, 2019, president Donald Trump accused doctors of executing babies who would die soon after birth due to fatal anomalies after a failed abortion attempt.^26^ A family may choose palliative care or comfort care, that might involve “*swaddling the newborn in a blanket and allowing the baby to die naturally without medical intervention*”. It ignited national debates over abortion and palliative care for newborns on Twitter (e.g., tweets like “*Some infants are born with … anencephaly where they have no brain. They cannot survive and need palliative, compassionate end of life care. Trump has no Compassion! So of course, he would let babies live in pain for days!*”. We observed that many of these consumer discussions happened after the spread of “*promotional information*” on these topics; and there are strong correlations on these topics between consumer and promotional tweets.

Further, the high-quality topics are similar between promotional information and consumer’s discussions, where 12 out of the 17 high-quality topics are the same across the two. We found that 9 out of the 12 common topics in consumer tweets are correlated (p < 0.05) with promotional information, suggesting that promotional health information on Twitter certainly has an impact on consumer discussions. This finding is consistent with our previous study on Lynch syndrome and breast cancer.^12^ The strong correlations might also indicate that promoting public awareness of palliative care through Twitter may be an effective health communication strategy. For RQ2, 16 high-quality topics in consumer palliative care discussions can be mapped to IBM constructs. Most of these topics are related to people’s attitude (9 topics) and perceived norm (5 topics). However, constructs such as personal agency, salience of the behavior, and habit are not found in these topics. One possible reason is that compared with attitude and perceived norm, constructs such as personal agency (i.e., individual’s capability to originate the behavior) are rarely discussed on Twitter. People are more willing to talk about their feelings and perceived norms (e.g., *“People ask for help and pray for hospice patients”*) than personal agency in performing the behavior. We then explored the correlation between the determinants measured from HINTS survey and learned topics from Twitter based on the geographic distributions. 14 out of the 17 high-quality consumer topics are correlated (p < 0.05) with HINTS responses, suggesting findings from social media data are comparable to—thus, may be an additional data source to supplement—those from traditional survey methods. One of the highest correlations we found is between topic 31/45: “*family members in hospice*” and QG2.b: Strongly/Somewhat disagree on “*To me, the goal of palliative care is to help friends and family to cope with a patient’s illness*” (i.e., *ρ* = -0.39, p = 0.02). In another word, in states where less people discuss palliative and hospice care (as observed in Twitter data), the more people disagree with the correct statements of those care (as observed in HINTS). A possible explanation is that in states with more discussions on family member in hospice will increase an individual’s perceived norm towards palliative care use, which will increase her positive feelings (i.e., attitude) towards the goal of palliative care. Through this example, social media findings can be an excellent supplementary data source to traditional methods providing additional and deeper insights. For RQ3, we successfully built models to accurately identify Twitter user’s gender based on her postings (i.e., the best performed CNN classifier has an F1-score of 0.8). Our CNN model identified 6,322 males and 20,787 females out of the 27,089 users who are considered as lay consumers. More female than male users joined palliative care-related discussions. Following the same approach as RQ2, we calculated the correlations between Twitter consumer topics and HINTS responses stratified by gender. Out of the 17 high-quality consumer topics, 8 common topics have strong correlations with HINTS responses in all 3 groups of analysis (i.e., consider “both gender”, “male”-only and “female”-only as examples shown in ***Table 5***), 6 topics have strong correlations in “both gender” and “female”-only groups, no topic in “both gender” and “male”-only groups, 2 topics in “female”-only and “male”-only groups, 1 topic in “male”-only group, and no topic in “female”-only group. Our findings indicate (1) strong gender differences when discussing palliative care-related topics, and (2) significant variations on how social media findings are correlated to survey responses when the gender variable can be considered. This suggests the importance of being able to tease out user attributes like gender in social media studies.

We also need to recognize the limitations of social media studies. First, beside the classifiers we used in this paper, there are many other the state-of-art algorithms (e.g., ensemble models). We did not exhaustively experimented with all methods. Second, social media users (i.e., Twitter users) are different from the general population (i.e., HINTS survey respondents). Thus, a strong correlation of between Twitter topics with HINTS responses does not indicate that findings from Twitter data can be translated or applied directly to the general population. Twitter users are younger than the general population. Thus, the representativeness of social media populations should be carefully considered when interpreting study findings. Third, there are lots of noise data in the Twitter, like ads bots and fake accounts, which may also distort the representativeness of our findings.

Our study demonstrated that social media like Twitter offer unique opportunities to assess consumers’ health behaviors. Nevertheless, social media data can only supplement traditional methods and biases of social media data need to be carefully considered when interpreting the study results.

## Data Availability

The data that support the findings of this study are openly available in https://github.com/yup111/twitter_palliative_care_data at github.com.

## Acknowledgements

This study was supported by the University of Florida Health Cancer Center (UFHCC) Research Pilot Grant and in part by NSF Award #1734134 and NIH UL1TR001427. The content is solely the responsibility of the authors and does not necessarily represent the official views of the sponsors.

